# Impact of COVID-19 pandemic and lockdown measures on mental health of children and adolescents in Greece

**DOI:** 10.1101/2020.10.18.20214643

**Authors:** Konstantina Magklara, Helen Lazaratou, Anastasia Barbouni, Konstantinos Poulas, Konstantinos Farsalinos, Coronavirus Greece Research Group

**Author notes:** Corresponding authors Konstantinos Poulas, Konstantinos Farsalinos. Authors equally contributed to the manuscript (Konstantina Magklara, Helen Lazaratou). Funding. No funding was provided for this study. All authors have seen and approved the manuscript.

## Abstract

**Background:** Mental health effects of the COVID-19 pandemic and the subsequent lockdown measures are expected to be profound.

**Aim:** The aim of the present study was to investigate the impact of the pandemic and the lockdown on children’s and adolescents’ mental health in Greece.

**Methods:** A cross-sectional survey of 1,232 Greek parents of children and adolescents aged < 18 years was conducted in March and May 2020. Parents provided information about sociodemographic characteristics, family everyday life during the lockdown and the pandemic psychological impact on their children.

**Results:** Approximately one-third (35.1%) of parents reported that the psychological health of their children was considerably affected. The most significant concern was social isolation. Unemployment, increased family conflicts, no opportunity for tele-work and a deteriorating psychological health of the parent, as well as children’s previous history of physical health conditions were all significantly associated with adverse mental health impact.

**Conclusion:** A considerable proportion of children and adolescents may experience adverse mental health effects due to the COVID-19 pandemic and the lockdown measures, and socioeconomic inequalities may be associated with these effects.

## Introduction

During the last few months children and adolescents around the world have experienced a major disruption of their everyday lives and daily routines due to the COVID-19 pandemic. In Greece, as in many other countries, regulatory measures have been introduced in early spring, in order to mitigate the risk of SARS-CoV-2 virus transmission. On March 10, all schools and universities closed and reopened eight to ten weeks later. All sport and leisure activities were suspended, while about 80% of private sector services providing psychological treatments for children and adolescents with mental health problems (reimbursed by the National Organization for the Provision of Health Services) closed.^1^

During the first wave of the pandemic in spring, Greece managed to contain the pandemic and had a relatively low number of confirmed COVID-19 cases and deaths compared to other countries.^2^ However the type, duration and intensity of the implemented measures had not been different from those imposed by other countries experiencing a less favorable course of the outbreak. Additionally, the Greek health system has only recently started recovering from a fiscal crisis with profound socioeconomic implications. During the past years there were significant cuts in hospital budgets, understaffing, occasional shortages of medical supplies and limited access to care and preventive services.^3^ Despite increased pressure on mental health services during public health crises,^4-6^ these services usually suffer major cuts as they often lack a strong advocacy base to support their necessity, contrary to services targeting physical health. Pediatric mental health services seem to be particularly vulnerable to austerity measures. In Greece, child and adolescent mental health services and supportive policies have undergone major budget changes. Public funding cuts led to some services not being fully operational, while many non-profit child and adolescent mental health community centers, psychosocial rehabilitation units, and highly specialized establishments were suspended,^7^ while the number of abused or neglected children admitted for child protection to pediatric hospitals increased dramatically.^8^

The mental health effects of the COVID-19 pandemic and the subsequent lockdown measures are expected to be profound and possibly long-lasting, especially among the most vulnerable. It is anticipated that children and adolescents are going to be highly affected.^9-17^ Large-scale disastrous events are almost always accompanied by increases in posttraumatic stress disorder (PTSD), depression, anxiety, substance use disorder, sleep disturbances, various types of other mental and behavioral disorders, domestic violence and child abuse.^18^ The mental health impact of such events may occur in the immediate aftermath of the event and then persist over longer periods. For instance, the SARS epidemic in 2003 was associated with increases in PTSD, anxiety and general psychological distress in both patients and healthcare workers^19^. Relevant literature investigating the type of psychopathology after large-scale events in the population of children and adolescents is relatively scarce.^20-22^ Published data from China, which was the first country affected by the new coronavirus, show that clinging, inattention and irritability were the most prevalent psychological conditions demonstrated during the COVID-19 outbreak by children and adolescents of all age groups.^23^ The CoRonavIruS health and Impact Survey (CRISIS), which used a robust questionnaire covering key domains relevant to mental distress and resilience during the pandemic, showed that pre-existing mood states, perceived COVID risk, and lifestyle changes are strongly associated with negative mood states during the pandemic in population samples of adults and in parents reporting on their children in the US and UK.^24^ A recent rapid systematic review investigating the impact of social isolation and loneliness on the mental health of children and adolescents demonstrated an increased risk of depression and possibly anxiety, both at the time loneliness was measured and between 0.25 to 9 years later.^25^

The aim of the present study was to investigate the impact of the COVID-19 pandemic and the measure of lockdown on the mental health of children and adolescents in Greece. More specifically, the study investigated the type of symptomatology presented by a sample of children and adolescents in Greece during the course of the pandemic, as reported by their parents, as well as its association with socioeconomic factors.

## Methods

The study was a cross-sectional online survey of a convenience sample of Greek parents of children aged < 18 years. A questionnaire was designed and was uploaded in an online survey tool (www.surveymonkey.com). Initially, participants were presented with an informed consent, consisting of a brief presentation of the research group and the purpose of the study. They were asked to read and provide their consent before proceeding to the questionnaire. If they agreed, they were forwarded to the questions of the survey. Our survey took place between April and May 2020.

The questionnaire consisted of three sections. The first section included general sociodemographic variables for parents and their offspring (parental sex, age, nationality, geographical area of current address, type of residential area, highest educational level attained, and employment status; child’s sex and age), as well as information regarding family and/or household characteristics (family annual income, number of bedrooms, number of household members). Information relevant to the pandemic such as parental tele-working and employment as a healthcare worker during the outbreak were also sought. The second section included questions about conditions of the everyday life of the family during the outbreak period. We obtained information about conflicts within the family the level of contact with friends and relatives and self-perceived psychological health of parents. The third section included information about the psychological impact of the pandemic on children. We asked about the child’s psychological health (current and before the outbreak), current psychological symptoms (sleep, stress, fear of being infected with the coronavirus, negative thoughts, depressive mood), physical activity and nutrition. We developed the questionnaire for the needs of the current study based on the tools used by “The CoRonavIruS Health Impact Survey” (CRISIS).^24^

To improve the design of the data collection instrument and to avoid potential comprehension issues experienced by the study participants, 8 parents were recruited to pretest the questionnaire using the method of cognitive interviewing.^26^ Once finalized and released online, the study and the survey instrument was advertised in social media while a press release was distributed to journalists who presented the study inviting participants in internet news-media. The study was approved by the ethics committee of the University of West Attica in Athens, Greece. Survey participation was anonymous and participants were informed through the consent form that they could exit the questionnaire at any time. The IP addresses were recorded by the online system in order to prevent double entries.

### Statistical analysis

Results were reported for the whole sample, with categorical variables presented as number (%) and continuous variables as mean (SD). Chi-square tests were used to compare characteristics between those with and without a negative impact of the pandemic on children’s psychological health. Logistic regression analysis was performed to examine factors associated with an impact of the pandemic on children’s psychological health. All analyses were performed using SPSS v.22 (Chicago, IL).

## Results

The sample of our study consisted of 1,232 parents who completed the online questionnaire. **Table 1** shows the basic sociodemographic and socioeconomic characteristics of participants. Most of them were of Greek nationality (98.2%), middle-aged women (79.6%), highly educated (90.0%) and residing in big cities (81.3%). Approximately half of the unemployed parents (7.0% out of a total unemployment rate of 15.0%) reported that they were employed before the pandemic, but lost their jobs during the lockdown. The vast majority of participants reported a family annual income of ≥ 10,000 euros, while in more than half of the households (55.9%) had more than 23 bedrooms in their home. Most of the participants were parents of school-aged children (70.4%), with almost 1 out of 5 (19.5%) reporting their current psychological health as fair or bad. A positive history of mental or developmental disorder was reported for 5.1% of the children, while a history of physical health condition was reported by 5.5%.

**Table 1.**
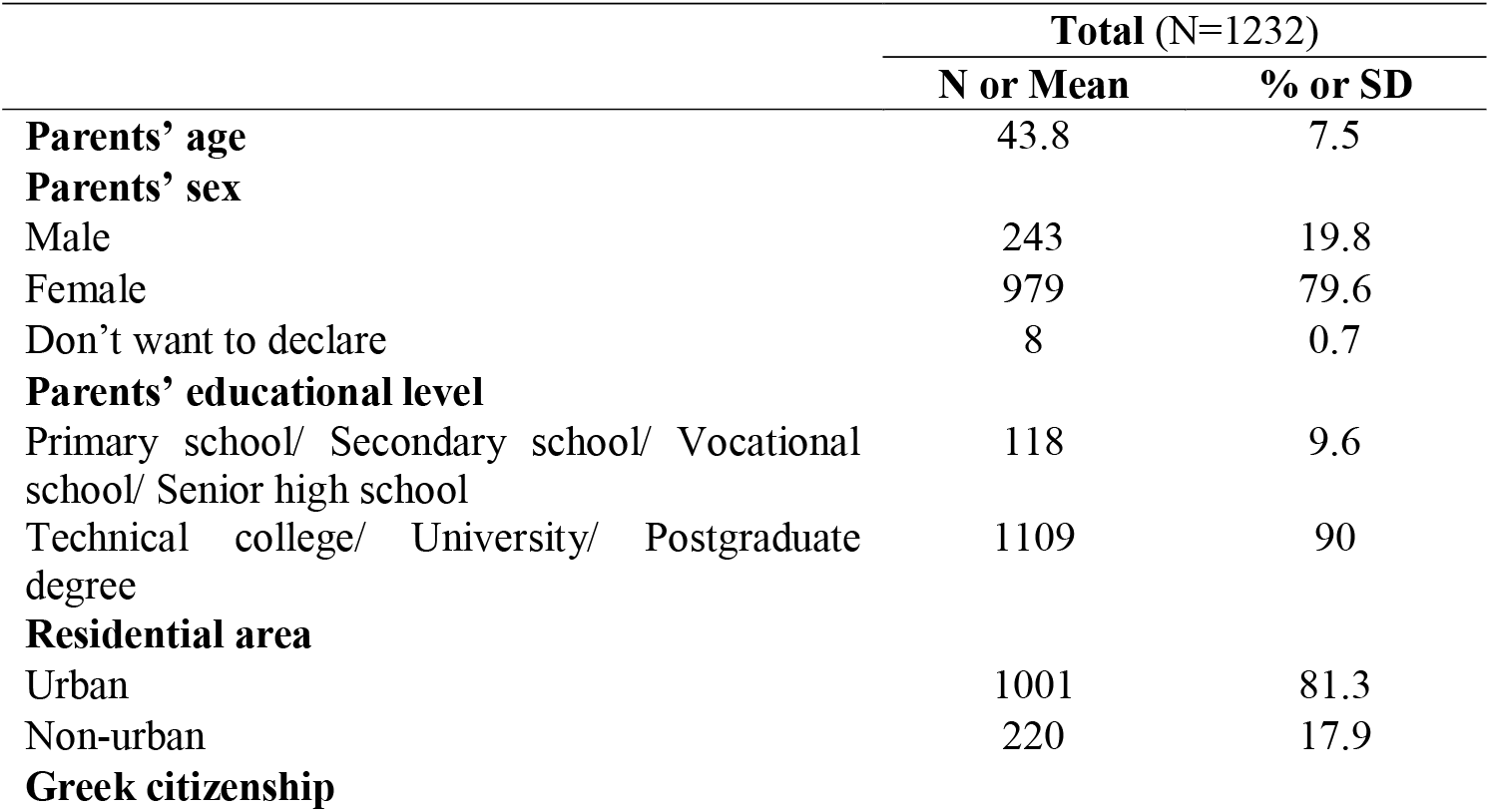
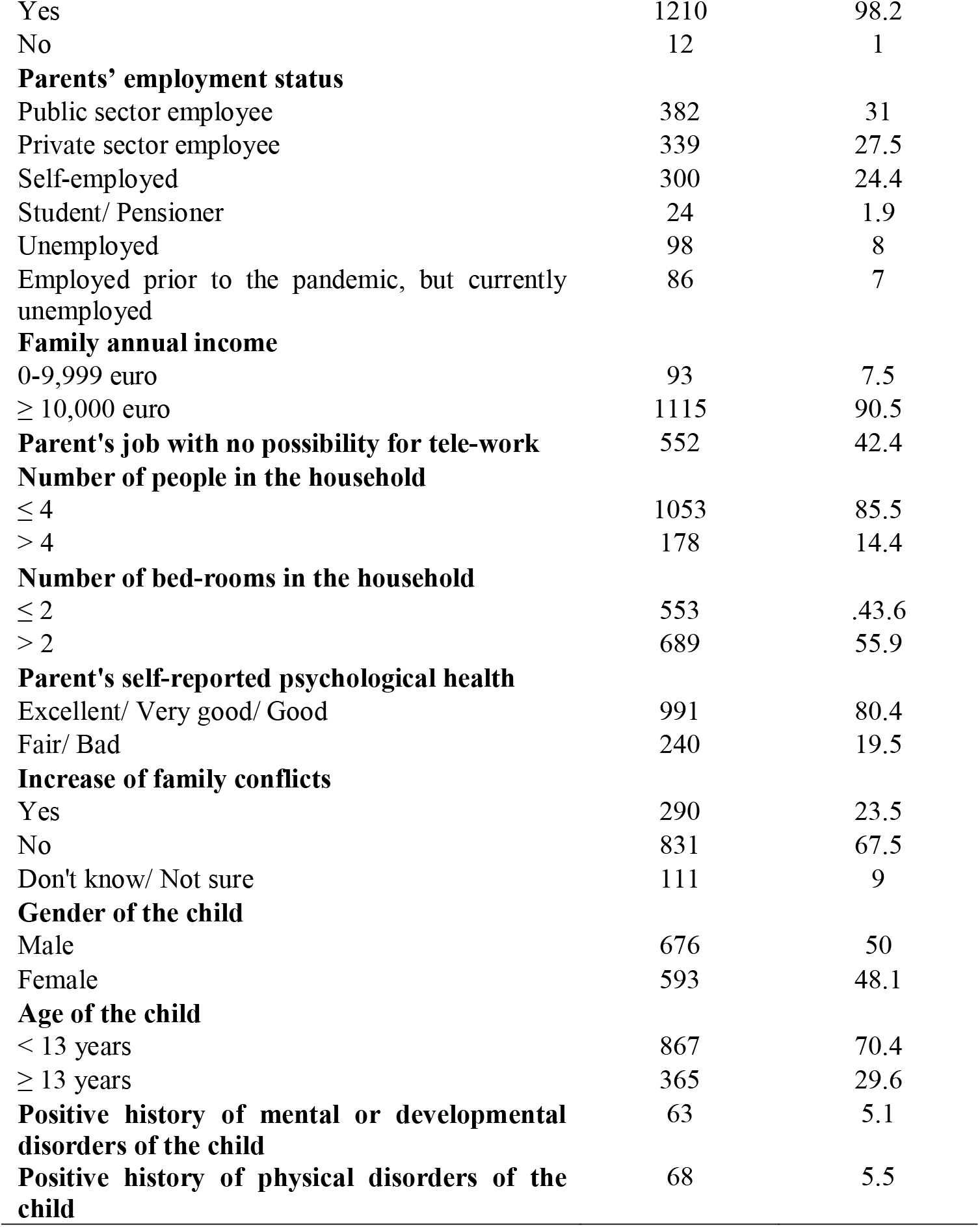
Basic sociodemographic and socioeconomic characteristics in a sample of 1,232 parents in Greece.

|  | Total (N=1232) |  |
| --- | --- | --- |
|  | N or Mean | % or SD |
| **Parents' age** | 43.8 | 7.5 |
| **Parents' sex** |  |  |
| Male | 243 | 19.8 |
| Female | 979 | 79.6 |
| Don't want to declare | 8 | 0.7 |
| **Parents' educational level** |  |  |
| Primary school/ Secondary school/ Vocational school/ Senior high school | 118 | 9.6 |
| Technical college/ University/ Postgraduate degree | 1109 | 90 |
| **Residential area** |  |  |
| Urban | 1001 | 81.3 |
| Non-urban | 220 | 17.9 |
| **Greek citizenship** |  |  |
| Yes | 1210 | 98.2 |
| No | 12 | 1 |
| **Parents' employment status** |  |  |
| Public sector employee | 382 | 31 |
| Private sector employee | 339 | 27.5 |
| Self-employed | 300 | 24.4 |
| Student/ Pensioner | 24 | 1.9 |
| Unemployed | 98 | 8 |
| Employed prior to the pandemic, but currently unemployed | 86 | 7 |
| **Family annual income** |  |  |
| 0-9,999 euro | 93 | 7.5 |
| ≥ 10,000 euro | 1115 | 90.5 |
| **Parent's job with no possibility for tele-work** | 552 | 42.4 |
| **Number of people in the household** |  |  |
| ≤ 4 | 1053 | 85.5 |
| > 4 | 178 | 14.4 |
| **Number of bed-rooms in the household** |  |  |
| ≤ 2 | 553 | 43.6 |
| > 2 | 689 | 55.9 |
| **Parent's self-reported psychological health** |  |  |
| Excellent/ Very good/ Good | 991 | 80.4 |
| Fair/ Bad | 240 | 19.5 |
| **Increase of family conflicts** |  |  |
| Yes | 290 | 23.5 |
| No | 831 | 67.5 |
| Don't know/ Not sure | 111 | 9 |
| **Gender of the child** |  |  |
| Male | 676 | 50 |
| Female | 593 | 48.1 |
| **Age of the child** |  |  |
| < 13 years | 867 | 70.4 |
| ≥ 13 years | 365 | 29.6 |
| **Positive history of mental or developmental disorders of the child** | 63 | 5.1 |
| **Positive history of physical disorders of the child** | 68 | 5.5 |

**Figure 1** shows children’s difficulties associated with the lockdown as reported by the parent. The parents in our sample reported that social isolation and the increase in screen time were the most significant difficulties experienced by the child or adolescent during the lockdown, followed by the decrease in physical activity and absence from educational activities.

**Figure 1.**
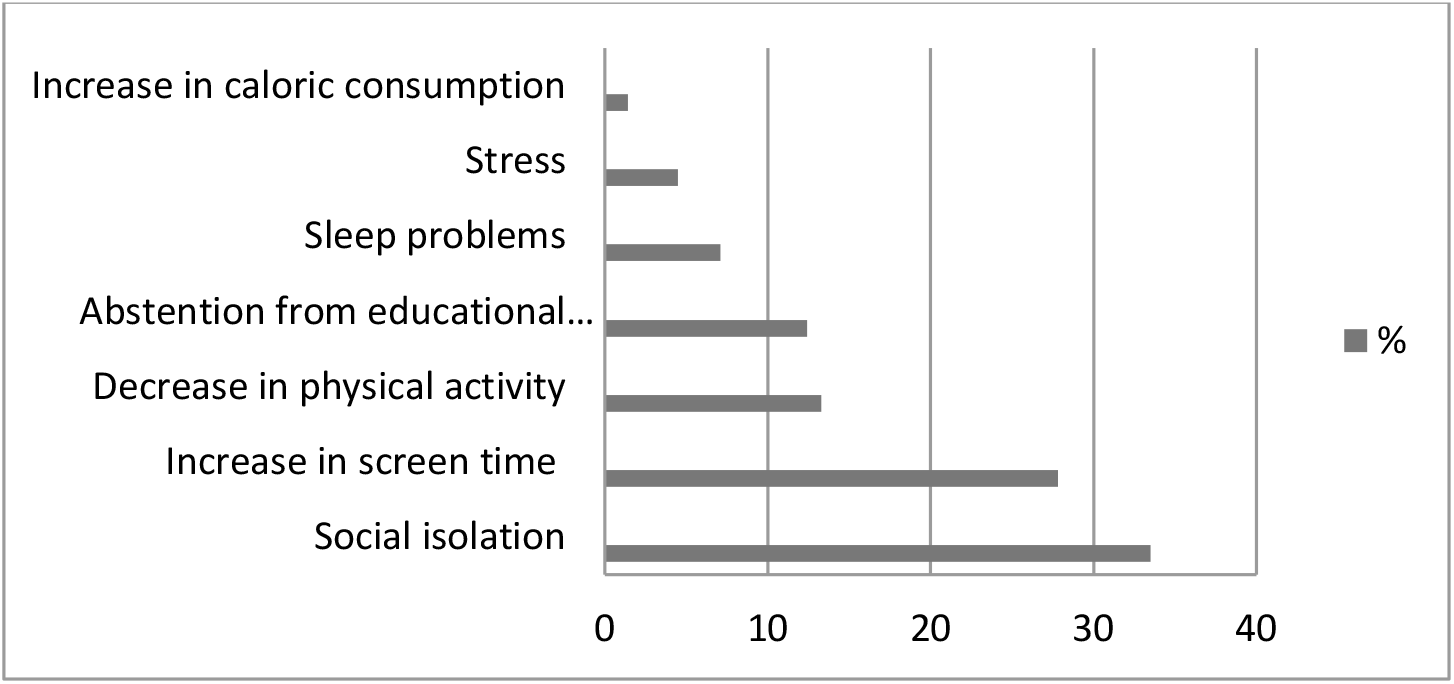
Difficulties experienced by children and adolescents during the lockdown as reported by the parent in a sample of 1,232 parents in Greece.

Approximately one-third of participants reported that the psychological health of their child was negatively affected by the lockdown. **Table 2** presents the characteristics of these cases compared to those with no impact of the lockdown on child’s psychological health. Statistically significant differences between the two groups were observed in the employment status of the parent, the opportunity for the parent to work from home, the increase in family conflicts, parent’s psychological condition and the positive history of mental or developmental and physical health condition of the child.

**Table 2.**
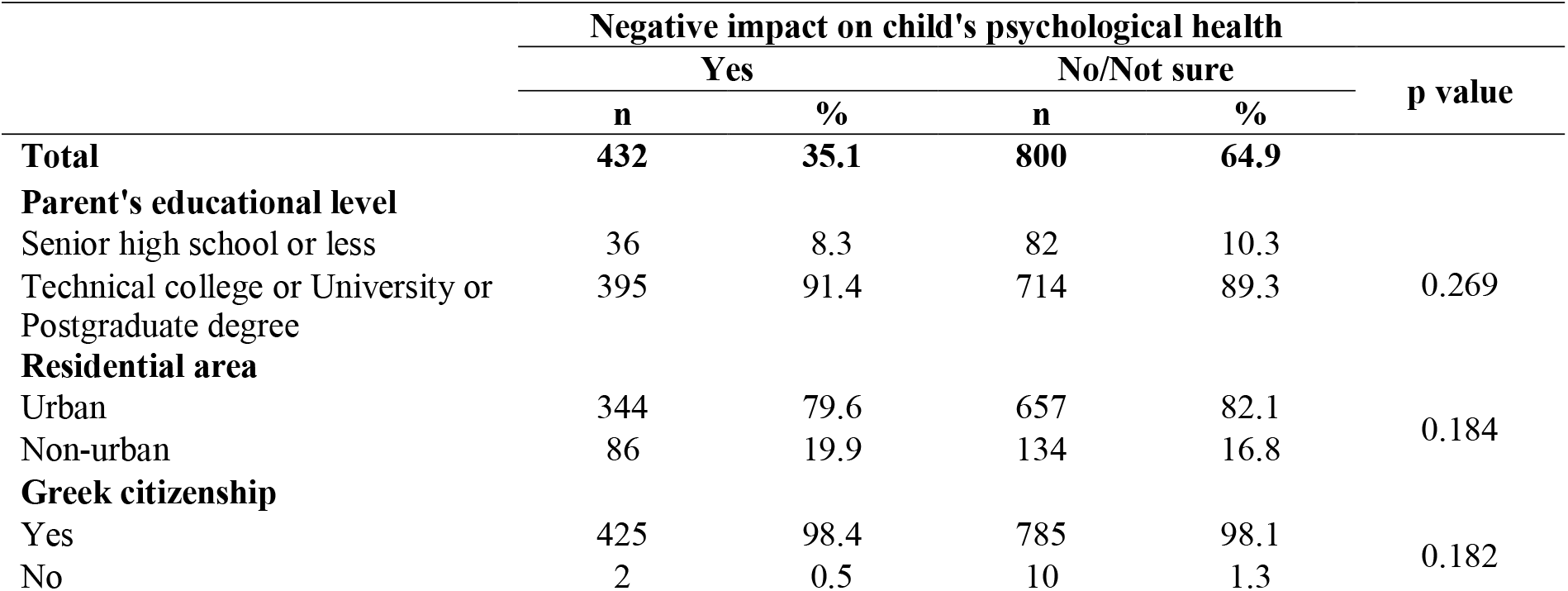
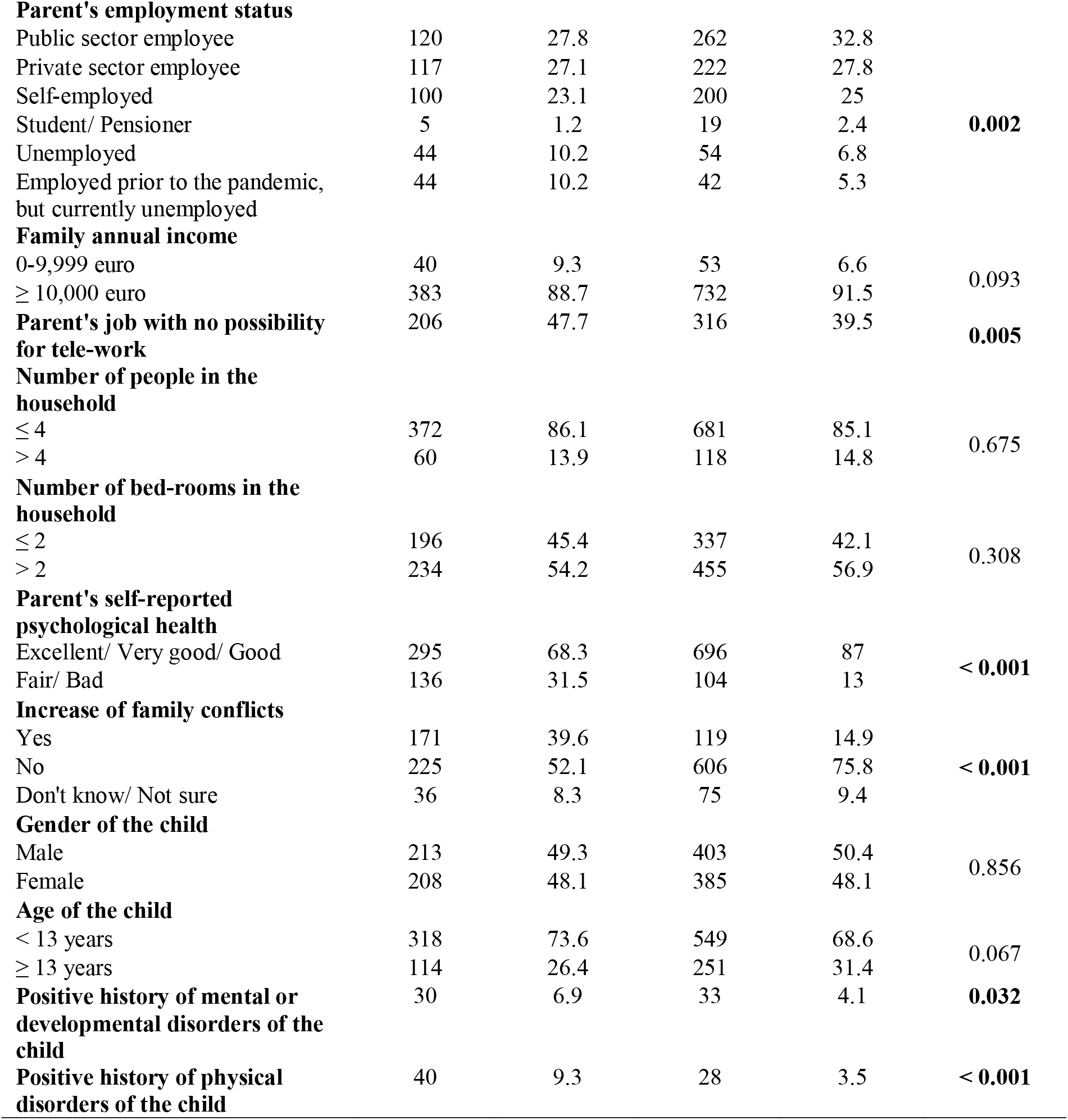
Characteristics of children without and with a negative impact of the pandemic on their psychological health as reported by the parent in a sample of 1,232 parents in Greece.

**Table 3** presents the results of logistic regression analysis. Unemployment of the parent, both previously existing and having occurred during the lockdown, parent having no opportunity to work from home, increase in family conflicts, impaired psychological condition of the parent during the lockdown and child’s previous history of a physical health condition were all significantly associated with a higher risk of negative impact of the lockdown on the child’s psychological health.

**Table 3.**
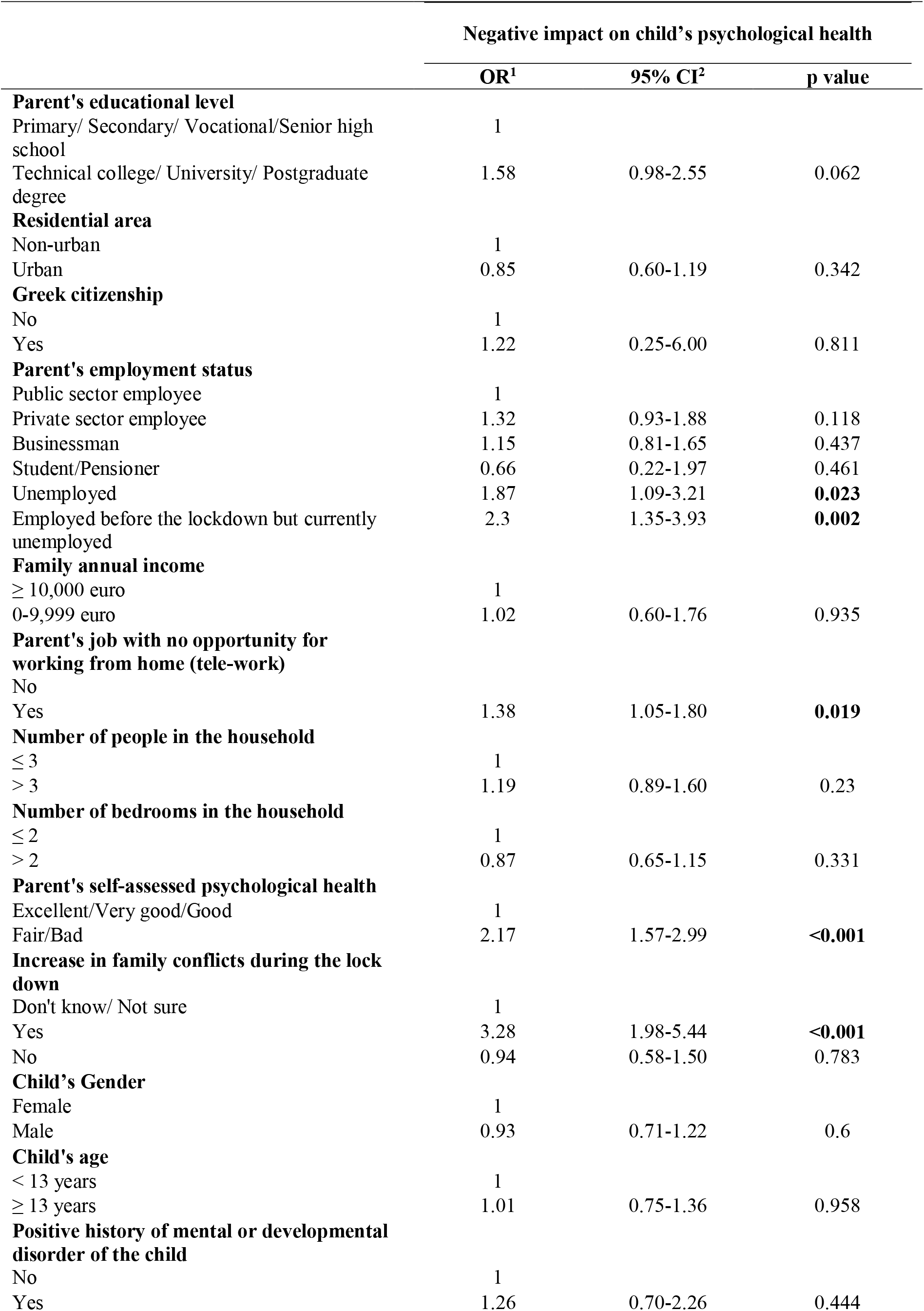
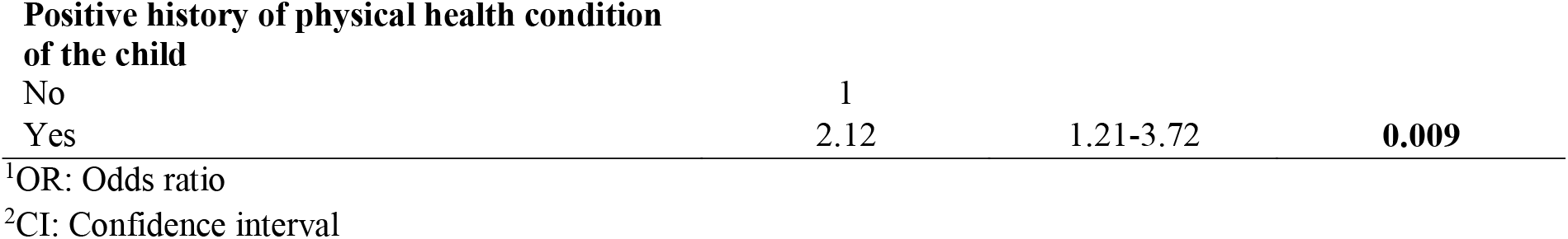
Logistic regression analysis of factors associated with an impact of the pandemic on children’s psychological health in a sample of 1,232 parents in Greece.

|  | Negative impact on child's psychological health |  |  |
| --- | --- | --- | --- |
|  | OR<sup>1</sup> | 95% CI<sup>2</sup> | p value |
| **Parent's educational level** |  |  |  |
| Primary/ Secondary/ Vocational/Senior high school | 1 |  |  |
| Technical college/ University/ Postgraduate degree | 1.58 | 0.98-2.55 | 0.062 |
| **Residential area** |  |  |  |
| Non-urban | 1 |  |  |
| Urban | 0.85 | 0.60-1.19 | 0.342 |
| **Greek citizenship** |  |  |  |
| No | 1 |  |  |
| Yes | 1.22 | 0.25-6.00 | 0.811 |
| **Parent's employment status** |  |  |  |
| Public sector employee | 1 |  |  |
| Private sector employee | 1.32 | 0.93-1.88 | 0.118 |
| Businessman | 1.15 | 0.81-1.65 | 0.437 |
| Student/Pensioner | 0.66 | 0.22-1.97 | 0.461 |
| Unemployed | 1.87 | 1.09-3.21 | **0.023** |
| Employed before the lockdown but currently unemployed | 2.3 | 1.35-3.93 | **0.002** |
| **Family annual income** |  |  |  |
| ≥ 10,000 euro | 1 |  |  |
| 0-9,999 euro | 1.02 | 0.60-1.76 | 0.935 |
| **Parent's job with no opportunity for working from home (tele-work)** |  |  |  |
| No | 1 |  |  |
| Yes | 1.38 | 1.05-1.80 | **0.019** |
| **Number of people in the household** |  |  |  |
| ≤ 3 | 1 |  |  |
| > 3 | 1.19 | 0.89-1.60 | 0.23 |
| **Number of bedrooms in the household** |  |  |  |
| ≤ 2 | 1 |  |  |
| > 2 | 0.87 | 0.65-1.15 | 0.331 |
| **Parent's self-assessed psychological health** |  |  |  |
| Excellent/Very good/Good | 1 |  |  |
| Fair/Bad | 2.17 | 1.57-2.99 | **<0.001** |
| **Increase in family conflicts during the lockdown** |  |  |  |
| Don't know/ Not sure | 1 |  |  |
| Yes | 3.28 | 1.98-5.44 | **<0.001** |
| No | 0.94 | 0.58-1.50 | 0.783 |
| **Child's Gender** |  |  |  |
| Female | 1 |  |  |
| Male | 0.93 | 0.71-1.22 | 0.6 |
| **Child's age** |  |  |  |
| < 13 years | 1 |  |  |
| ≥ 13 years | 1.01 | 0.75-1.36 | 0.958 |
| **Positive history of mental or developmental disorder of the child** |  |  |  |
| No | 1 |  |  |
| Yes | 1.26 | 0.70-2.26 | 0.444 |
| **Positive history of physical health condition of the child** |  |  |  |
| No | 1 |  |  |
| Yes | 2.12 | 1.21-3.72 | **0.009** |

<sup>1</sup>OR: Odds ratio
<sup>2</sup>CI: Confidence interval

## Discussion

One out of three parents in a sample of 1,232 parents in Greece reported that the most significant difficulty experienced by their offspring during the lockdown was social isolation. Other important issues were the increase in screen time and the decrease in physical activity, as well as the absence from educational activities. Approximately one-third of the parents reported that the psychological health of their children was considerably affected during the crisis. The unemployment of the parent and their deprivation from opportunities for tele-work was significantly associated with the impact of the lockdown on the mental health of their offspring. Increased family conflicts and a deteriorating psychological health of the parent during the lockdown correlated also with the psychological health of children and adolescents. Previous history of a physical health condition of the child was statistically significantly associated with a higher risk of an impact of the measures on the psychological health of the child. There was a trend for a similar association with previous history of a mental or developmental disorder of the child, which however did not reach the level of statistical significance.

One of the major concerns of the parents in our sample during the lockdown was the social isolation of their children. In many countries around the world children and adolescents have experienced an unexpected and prolonged period of physical isolation from their friends, peers, teachers, extended family and various community networks. Literature shows that isolation associated with quarantine measures has negative psychological effects in adults, including confusion, anger and post-traumatic distress.^11^ Loneliness might be a major issue associated with social isolation. Loneliness is conceptualized as the negative emotion of a discrepancy between the social contact actually experienced by a person and the desire for it.^27^ Social isolation is not necessarily followed by loneliness. However, early evidence shows that more than one third of adolescents report high levels of loneliness,^28^ while a rapid systematic review of previous literature reports that social isolation and loneliness increases the risk of depression and possibly anxiety.^25^

Further concerns of the parents in our sample were the limited physical activity and the increased screen time of children and adolescents during the lockdown. A study conducted in 2,426 children and adolescents in Shanghai, China confirmed a substantial decrease in physical activity and a significant increase in screen time during the COVID-19 pandemic.^29^ The vicious circle between reduced physical activity and increased sedentary behavior was one of the major concerns early in the crisis that lead in an urgent call for systematic efforts to mitigate the effects of home confinement of children.^17,30^

Our study presents some interesting findings suggesting socioeconomic inequality in the distribution of the negative effects of the COVID-19 crisis in the population of children and adolescents. The unemployment of the parent that was either present prior or occurred during the course of the pandemic was significantly associated with a negative impact on children’s psychological health. Socioeconomic inequalities are among the most common findings in literature investigating the mental health of children and adolescents.^31,32^ There is evidence that higher income inequality relates not only to more psychological and physical symptoms, but also to fewer days of physical activity and higher Body Mass Index, which is in accordance with the higher concern of reduced physical activity reported in our study.^31^ Parental unemployment is one of the most significant socioeconomic indicators associated with the psychological health of children and adolescents. Financial stress is considered to be the most important consequence of unemployment, with regard to the health of the unemployed individual and the family members.^32,33^ The significance of the above-mentioned associations is of particular interest for Greece, since it is a country that was severely affected by a major fiscal crisis during the last decade.^3^ According to a report conducted on behalf of UNICEF three years ago, almost one in two children in Greece (45%) lived in material deprivation and 22% in severe material deprivation.^34^

Increased family conflicts and poorer psychological health of the parent have been also associated with higher risk of a negative impact of the crisis on the psychological health of children and adolescents in our study. Family conflict, in the form of either inter-adult or parent–child conflict, has been linked in various studies to children’s behavioral, emotional, social, academic and health problems and identified as important predictor of children’s maladjustment.^35^ Regarding the impact of parental psychological health, most existing findings derive from studies conducted in clinical populations, linking parental psychopathology to a wide range of negative outcomes for their offspring.^36^ Population studies have shown similar results reporting positive associations between parental psychological distress and psychosocial maladjustment of their offspring.^37,38^

Finally, our study showed a positive association between a history of a physical health condition and a negative impact of the lockdown on the psychological health of the child. Chronic physical illness is thought to be a risk factor for psychological problems. The presence of physical symptoms and the need for disease management strategies are likely to interfere with many aspects of the child’s everyday life, may cause frustration and detrimentally affect the child’s self-esteem. Additionally, chronic illness may lead to problematic parental behaviors, ranging from overprotection to rejection, which may further impair the child’s psychological health.^39^ There was evidence of a trend for a similar association with a history of previous psychiatric or developmental disorder, which has not reached however the level of statistical significance in our sample. Multiple studies point out a possibly higher vulnerability of individuals with preexisting mental illness that experience additional adverse events.^24,40^ Thus it could be argued that in our sample the trend has not reached the level of statistical significance due to lack of statistical power.

### Limitations

An important limitation of our study is the fact that we used an online, convenience sample, which is not representative of the population studied. It has been argued that non-representative samples attract volunteers who are already well engaged, interested in the topic, who can access the internet, while those excluded, for instance individuals with severe mental illness, are often those most in need.^41^ Therefore, our findings cannot be interpreted as an overall prevalence of psychopathology among children in the general population. However, our results indicate that individuals with various types of vulnerabilities are at higher risk for negative outcomes of the current crisis. Given the argument that vulnerable populations may have been excluded from our surveys, it could be expected that the real impact of COVID-19 pandemic could be of greater magnitude for specific population sub-groups.

Another limitation of our study is the fact that we used only parental reports. However, we collected our data during the lockdown and there were only limited opportunities to use more subjective measures of children’s characteristics and difficulties. Additionally, the online nature of the design of our study presented significant difficulties for the use of children as informants, especially for younger children that were the majority in our sample.

## Conclusions

Our study shows that vulnerable individuals are at higher risk for experiencing negative outcomes during the course, and possibly also at the aftermath, of the COVID-19 pandemic. Both family risk factors, such as parental unemployment, psychological distress or family conflict, and individual risk factors, such as preexisting physical health conditions, contribute substantially to the vulnerability of children and adolescents to the adverse effects of the new coronavirus outbreak and its containment measures. Well-designed public health policies should focus on specific population groups with the aim of ameliorating risks, when possible, and enhancing resilience.

## Data Availability

The dataset will be available upon reasonable request.

## References

1. Giannopoulou, I., & Tsobanoglou, G. O. (2020). COVID-19 pandemic: challenges and opportunities for the Greek health care system. Irish Journal of Psychological Medicine, 1–9.

2. Johns Hopkins Center for Systems Science and Engineering, https://systems.jhu.edu/research/public-health/ncov/. Date accessed: 28 September 2020

3. Kentikelenis, A., Karanikolos, M., Papanicolas, I., Basu, S., McKee, M., & Stuckler, D. (2011). Health effects of financial crisis: omens of a Greek tragedy. The Lancet, 378(9801), 1457–1458.

4. Durkheim, E. (2006). Durkheim: Essays on morals and education (Vol. 1). Taylor & Francis.

5. Frasquilho, D., Matos, M. G., Salonna, F., Guerreiro, D., Storti, C. C., Gaspar, T., & Caldas-de-Almeida, J. M. (2015). Mental health outcomes in times of economic recession: a systematic literature review. BMC public health, 16(1), 1–40

6. Weaver, J. D. (1983). Economic recession and increases in mental health emergencies. The journal of mental health administration, 10(2), 28–31.

7. Kolaitis, G., & Giannakopoulos, G. (2015). Greek financial crisis and child mental health. The Lancet, 386(9991), 335.

8. Koliatis 2014

9. Araújo, F. J. de O., de Lima, L. S. A., Cidade, P. I. M., Nobre, C. B. & Neto, M. L. R. Impact Of Sars-Cov-2 And Its Reverberation In Global Higher Education And Mental Health. Psychiatry Res. 288, 112977 (2020).

10. Asmundson, G. J., & Taylor, S. (2020). How health anxiety influences responses to viral outbreaks like COVID-19: What all decision-makers, health authorities, and health care professionals need to know. Journal of Anxiety Disorders, 71, 102211.

11. Brooks, S. K., Webster, R. K., Smith, L. E., Woodland, L., Wessely, S., Greenberg, N., & Rubin, G. J. (2020). The psychological impact of quarantine and how to reduce it: rapid review of the evidence. The Lancet.

12. Liu, C. H., Zhang, E., Wong, G. T. F., & Hyun, S. (2020). Factors associated with depression, anxiety, and PTSD symptomatology during the COVID-19 pandemic: Clinical implications for US young adult mental health. Psychiatry Research, 113172.

13. Luo, M., Guo, L., Yu, M. & Wang, H. The Psychological and Mental Impact of Coronavirus Disease 2019 (COVID-19) on Medical Staff and General Public – A Systematic Review and Meta-analysis. Psychiatry Res. 113190 (2020)

14. Mactavish, A., Mastronardi, C., Menna, R., Babb, K. A., Battaglia, M., Amstadter, A. B., & Rappaport, L. (2020). The Acute Impact of the COVID-19 Pandemic on Children’s Mental Health in Southwestern Ontario. Preprint, 19 September 2020

15. Mazza, M., Marano, G., Lai, C., Janiri, L., & Sani, G. (2020). Danger in danger: Interpersonal violence during COVID-19 quarantine. Psychiatry research, 113046.

16. Rogers, J. P., Chesney, E., Oliver, D., Pollak, T. A., McGuire, P., Fusar-Poli, P., … & David, A. S. (2020). Psychiatric and neuropsychiatric presentations associated with severe coronavirus infections: a systematic review and meta-analysis with comparison to the COVID-19 pandemic. The Lancet Psychiatry.

17. Wang, G., Zhang, Y., Zhao, J., Zhang, J., & Jiang, F. (2020). Mitigate the effects of home confinement on children during the COVID-19 outbreak. The Lancet, 395(10228), 945–947.

18. Neria Y, Nandi A, Galea S. (2008). Post-traumatic stress disorder following disasters: a systematic review. Psychol Med.; 38(4):467–480. doi:10.1017/S0033291707001353

19. Lee AM, Wong JG, McAlonan GM, et al. (2007) Stress and psychological distress among SARS survivors 1 year after the outbreak. Can J Psychiatry; 52(4):233–240. doi:10.1177/070674370705200405

20. Klein T.P., Devoe E.R., Miranda-Julian C., Linas K. (2009) Young children’s responses to September 11th: the New York City experience. Infant Ment Health J.;30:1–22

21. Hoven C.W., Duarte C.S., Lucas C.P., Wu P., Mandell D.J., Goodwin R.D. (2005) Psychopathology among New York City public school children 6 months after September 11. Arch Gen Psychiatry;62:545–552

22. Laor N., Wolmer L., Mayes L.C., Gershon A., Weizman R., Cohen D.J. (1997) Israeli preschool children under Scuds: a 30-month follow-up. J Am Acad Child Adolesc Psychiatry. 1997;36:349–356

23. Jiao, W. Y., Wang, L. N., Liu, J., Fang, S. F., Jiao, F. Y., Pettoello-Mantovani, M., & Somekh, E. (2020). Behavioral and emotional disorders in children during the COVID-19 epidemic. The journal of Pediatrics, 221, 264

24. Nikolaidis, A., Paksarian, D., Alexander, L., DeRosa, J., Dunn, J., Nielson, D. M., … & Milham, M. P. (2020). The Coronavirus Health and Impact Survey (CRISIS) reveals reproducible correlates of pandemic-related mood states across the Atlantic. medRxiv.

25. Loades, M. E., Chatburn, E., Higson-Sweeney, N., Reynolds, S., Shafran, R., Brigden, A., … & Crawley, E. (2020). Rapid Systematic Review: The Impact of Social Isolation and Loneliness on the Mental Health of Children and Adolescents in the Context of COVID-19. Journal of the American Academy of Child & Adolescent Psychiatry. https://doi.org/None.

26. Collins D. Pretesting survey instruments: an overview of cognitive methods. Qual Life Res. 2003 May;12(3):229–38. doi: 10.1023/a:1023254226592.

27. Perlman, D., & Peplau, L. A. (1981). Toward a social psychology of loneliness. Personal relationships, 3, 31–56.

28. VandePol, B. (2020). Resilience During COVID-19 on vimeo.html. https://archive.hshsl.umaryland.edu/handle/10713/12579 Date accessed: 28 September 2020.

29. Xiang, M., Zhang, Z., & Kuwahara, K. (2020). Impact of COVID-19 pandemic on children and adolescents’ lifestyle behavior larger than expected. Progress in Cardiovascular Diseases.

30. Guan, H., Okely, A. D., Aguilar-Farias, N., del Pozo Cruz, B., Draper, C. E., El Hamdouchi, A., … & Löf, M. (2020). Promoting healthy movement behaviours among children during the COVID-19 pandemic. The Lancet Child & Adolescent Health, 4(6), 416–418.

31. Elgar, F. J., Pförtner, T. K., Moor, I., De Clercq, B., Stevens, G. W., & Currie, C. (2015). Socioeconomic inequalities in adolescent health 2002–2010: a time-series analysis of 34 countries participating in the Health Behaviour in School-aged Children study. The Lancet, 385(9982), 2088–2095)

32. Magklara, K., Skapinakis, P., Niakas, D., Bellos, S., Zissi, A., Stylianidis, S., & Mavreas, V. (2010). Socioeconomic inequalities in general and psychological health among adolescents: a cross-sectional study in senior high schools in Greece. International journal for equity in health, 9(1), 3

33. Whelan, C. T. (1992). The role of income, life-style deprivation and financial strain in mediating the impact of unemployment on psychological distress: evidence from the Republic of Ireland. Journal of Occupational and Organizational Psychology

34. Papatheodorou C, Papathanasiou S. The state of the children in Greece report 2017: The children of the crisis. Hellenic National Committee for UNICEF, Athens, Greece 2017

35. Cummings, E. M., & Schatz, J. N. (2012). Family conflict, emotional security, and child development: translating research findings into a prevention program for community families. Clinical Child and Family Psychology Review, 15(1), 14–27

36. Zahn-Waxler C, Duggal S, Gruber R. Parental psychopathology. In: Bornstein MH, editor. Handbook of parenting: Vol. 4. Social conditions and applied parenting. 2. Mahwah, NJ: Erlbaum; 2002. pp. 295–327

37. Vostanis, P., Graves, A., Meltzer, H., Goodman, R., Jenkins, R., & Brugha, T. (2006). Relationship between parental psychopathology, parenting strategies and child mental health. Social psychiatry and psychiatric epidemiology, 41(7), 509–514

38. Roustit, C., Campoy, E., Chaix, B., & Chauvin, P. (2010). Exploring mediating factors in the association between parental psychological distress and psychosocial maladjustment in adolescence. European child & adolescent psychiatry, 19(7), 597–604

39. Bennett D S. Depression among children with chronic medical problems: A meta-analysis (1994), Journal of Pediatric Psychology, (19);149–169

40. Fiorillo, A., & Gorwood, P. (2020). The consequences of the COVID-19 pandemic on mental health and implications for clinical practice. European Psychiatry, 63(1).

41. Pierce, M., McManus, S., Jessop, C., John, A., Hotopf, M., Ford, T., … & Abel, K. M. (2020). Says who? The significance of sampling in mental health surveys during COVID-19. The Lancet Psychiatry.

